# Minimizing the infected peak utilizing a single lockdown: a technical result regarding equal peaks

**DOI:** 10.1101/2021.06.26.21259589

**Authors:** James M. Greene, Eduardo D. Sontag

**Affiliations:** Department of Mathematics, Clarkson University, Potsdam, NY, United States; Department of Electrical and Computer Engineering and Department of Bioengineering, Northeastern University, Boston, MA, United States; Laboratory of Systems Pharmacology, Program in Therapeutic Science, Harvard Medical School, Boston, MA, United States

## Abstract

Due to the usage of social distancing as a means to control the spread of the novel coronavirus disease COVID-19, there has been a large amount of research into the dynamics of epidemiological models with time-varying transmission rates. Such studies attempt to capture population responses to differing levels of social distancing, and are used for designing policies which both inhibit disease spread but also allow for limited economic activity. One common criterion utilized for the recent pandemic is the peak of the infected population, a measure of the strain placed upon the health care system; protocols which reduce this peak are commonly said to ‘flatten the curve.” In this work, we consider a very specialized distancing mandate, which consists of one period of fixed length of distancing, and addresses the question of optimal initiation time. We prove rigorously that this time is characterized by an equal peaks phenomenon: the optimal protocol will experience a rebound in the infected peak after distancing is relaxed, which is equal in size to the peak when distancing is commenced. In the case of a non-perfect lockdown (i.e. disease transmission is not completely suppressed), explicit formulas for the initiation time cannot be computed, but implicit relations are provided which can be pre-computed given the current state of the epidemic. Expected extensions to more general distancing policies are also hypothesized, which suggest designs for the optimal timing of non-overlapping lockdowns.

## 1 Introduction

The ongoing global COVID-19 (coronavirus disease 2019) pandemic, caused by SARS-CoV-2 (severe acute respiratory coronavirus 2), has necessitated the use of non-pharmaceutical interventions (NPIs) as a means to slow transmission of the disease. Although controversial, there is clear evidence that NPIs such as social distancing have saved millions of lives globally [1]. Social distancing mandates, denoted in this manuscript as “lockdowns,” cannot be implemented indefinitely, as it carries both a high economic [2, 3] and psychological [4] cost. Furthermore, a lack of compliance may make extended protocols unfeasible to implement [5, 6]. Hence there is a need to optimize the timing of prescribed lockdowns. The optimization of such schedules is the focus of this work. Specifically, we characterize the implementation time of a single *non-strict* lockdown, with fixed transmission reduction, which minimizes the peak of the infected population in the Susceptible-Recovered-Removed (SIR) model. The main contribution is Theorem 1, which we term an *equal peak* phenomenon.

There have been a large number of mathematical analyses applied to the spread of COVID-19. In this work, as we solve a small technical problem, we do not attempt to provide a comprehensive literature review. We do however note a number of closely related works based on optimal social distancing strategies. The analysis presented here is a direct extension of [7], where the case of multiple fixed-length non-overlapping *complete* (i.e. zero disease transmission) lockdowns is completely characterized as a linear programming problem; this manuscript should be viewed as a direct extension of this previous work. Again minimizing the infected peak, the authors of [8] determine the optimal (again, possibly complete) lockdown schedule as a feedback mechanism. Numerical results for a variety of epidemic objectives with respect to a single interval of distancing are provided in [9, 10], and [11] studies the same problem both numerically and theoretically. A constrained optimization problem is solved in [12], where the time minimal distancing policy which maintains an upper bounded on the infected population is derived. There are also a number of works which minimize the total number of infections during an epidemic during a period of such distancing; this is studied numerically in [9] and analytically in [13, 14, 15]. Interestingly, the main result of [13] is that the optimal lockdown policy to minimize the total number of infected individuals coincides with the protocol considered in this work; see equation (4) below. However, as observed in [9], the timing with respect to these differing objectives (minimizing infected peak vs. minimizing total number of infections) in general do *not* agree, so that policy makers cannot generally hope to achieve both simultaneously.

This work is organized as follows. In Section 2, we recall the SIR model and precisely formulate the social distancing protocol to be optimized. Results are presented in Section 3, and a discussion with potential (unproven) extensions are postulated in Section 4. Some preliminary numerical simulations are also provided in Section 4. Proofs of all results are provided in Section 5.

## 2 Problem formulation

We consider the classic SIR epidemic model introduced by Kermack and McKendrick in 1927 [16], which we briefly review here. The ordinary differential equations (ODEs) describing the evolution of the system are given below:

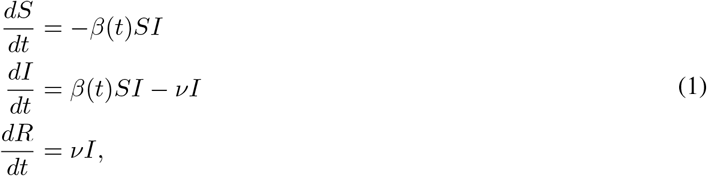

together with initial conditions

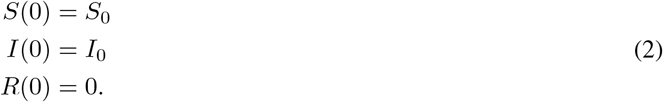

Here *S* denotes the susceptible population, *I* the infected population, and *R* the removed population, where the latter combines those individuals that have either obtained immunity or died. Parameter *ν* represents the combined recovery and death rate of the disease, and hence if mortality is relatively small, is a measure of the rate of recovery to immunity. We assume that *ν* is constant. Parameter *β* = *β*(*t*) quantifies the transmission rate between susceptible and infected individuals. We consider this a time-varying parameter, since NPIs are generally viewed as altering this transmission rate. Specifically, during a lockdown, where contacts are reduced and/or mask mandates are enforced, the transmission rate may be modeled as decreasing by a factor of *p*, where 0 ≤ *p* < 1:

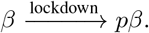

For example, in [17], estimated reductions in ℛ_0_ (which for the SIR model is equivalent to reducing *β*; see equation (7) below) yield *p* values as large as 0.58, with this value corresponding to mandates limiting gatherings to 10 people or less.

We are specifically interested in the effect of a single fixed period of social distancing (i.e. a single lockdown) on the peak of the infected population:

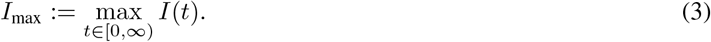

Our goal is thus to understand the behavior of *I*_max_ as a function of *β*(*t*), where *β*(*t*) take the following form:

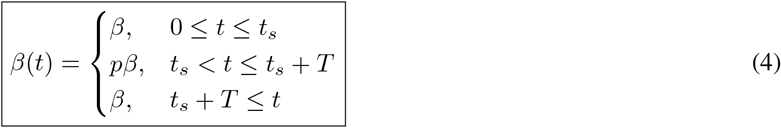

As discussed above, 0 ≤ *p* < 1 represents the reduction in transmission rate due to distancing mandates, which are enacted at time *t*_*s*_ for a length of time *T* . That is, a lockdown occurs for *t* ∈ [*t*_*s*_, *t*_*s*_ + *T*]. For a visualization of the lockdown protocol, see Figure 1. We assume that *β, ν, p*, and *T* are fixed and known, and we are interested in optimizing the start time *t*_*s*_ of distancing so as to minimize the infected peak as a function of *t*_*s*_:

**Figure 1:**
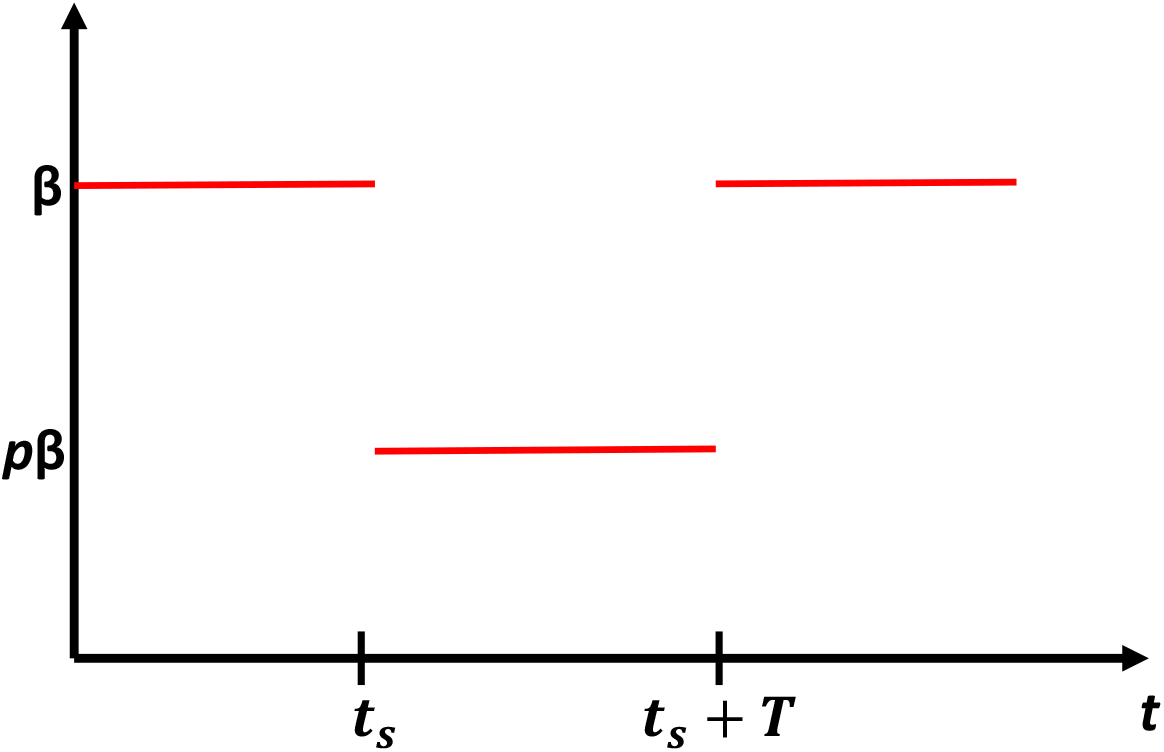
Visualization of idealized lockdown (4). We assume that transmission is reduced by an effective amount *p* during the lockdown, where 0 < *p* < 1. The lockdown is initiated at *t*_*s*_ and is enacted for *T* units of time.

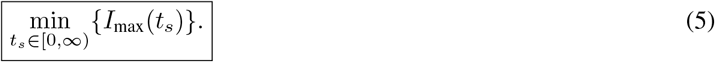

It is intuitively clear that *t*_∗_ should not occur too early or too late: begin too early and we simply delay the full effect of the epidemic, and begin too late and the epidemic has already passed throughout the susceptible population, and hence social distancing has minimal impact. It is the goal of this work to understand the optimal timing with respect to the metric (5), with transmission rate of the form (4). This problem is a generalization of [7], which studied the same problem for *p* = 0. In general we allow *p >* 0 in the following.

We lastly note the maximum appearing in (3) is indeed a maximum (i.e. it is achieved at some time, as opposed to a supremum), since *I* is continuous and *I*(*t*) →0 as *t*→∞ for all distancing protocols *β*(*t*). For more details and a proof, see for example [18].

We assume that the populations are normalized, so that

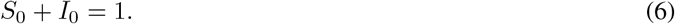

Assumption (6) ensures that *S*(*t*) + *I*(*t*) + *R*(*t*) = 1 for all *t* ≥ 0, i.e. that variables *S, I*, and *R* represent population fractions. System (1) is also seen to be positively invariant. Note lastly that the removed population *R* does not affect the dynamics of the above system, but may serve as a useful measure of disease progression.

### 2.1 Additional assumptions

In this section, we impose additional assumptions on the model introduced in Section 2. *We note that these assumptions are not crucial for the following theory, but instead are useful for limiting the number of potential cases which we feel would otherwise obfuscate the exposition*. Specifically, they allow us to conclude with certainty the exact locations of the potential relative maxima of the infected population curves, while excluding particular “boundary cases”.

Our first additional assumption is that the lockdown has the ability to immediately stop disease progression independently of the start time. In other words, we assume that the transmission rate is able to be reduced such that for any start time *t*_*s*_,

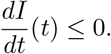

for all *t* ∈ [*t*_*s*_, *t*_*s*_ + *T*]. Since on this time interval

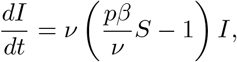

this assumption is equivalent to

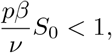

as *S* is non-increasing on [0, ∞). Defining the basic reproduction number ℛ_0_ as

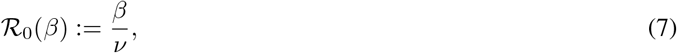

the previous assumption is equivalent to

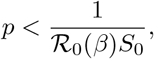

or again equivalently

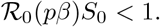

Furthermore, for an epidemic to exist (i.e. for *I* to increase at any time point, which is equivalent to increasing initially, since *S* is non-increasing), we must assume that *İ*(0) *>* 0. Examining the second of (1) (together with *β*(*t*) = *β* and the definition of ℛ_0_(*β*)), this implies that

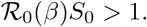

Thus, for the remainder of the manuscript, we assume the following:

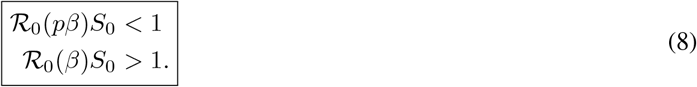

## 3 Results

### 3.1 Infected peak formulas

The assumptions presented in Section 2.1 imply that the graph of *I* : [0, ∞) → [0, 1] may have at most two local maxima, one of which must be global. In fact, there are exactly two possible cases, dependent on the lockdown initiation time *t*_*s*_:

1. *I* has a unique local maximum *I*_*max*_ occurring on [0, *t*_*s*_].
2. *I* has one maxima at *t*_*s*_ (*I*(*t*_*s*_)), and another local maximum on [*t*_*s*_ + *T*, ∞).

Note that Case 2 occurs precisely because of the second assumption in (8), i.e. since *I* is non-increasing on [*t*_*s*_, *t*_*s*_ + *T*]. More precisely, Case 2 occurs for *t*_*s*_ such that

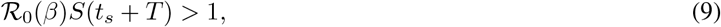

which in words means that *I* initially increases after the lockdown is released (i.e. at time *t*_*s*_ + *T*). Since *S* is non-increasing, (9) implies that

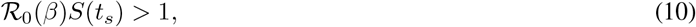

i.e. that *I* was also increasing prior to lockdown initiation.

It is not hard to see that the maximum in Case 1 is always larger than the pair of maxima in Case 2, so that the minimization problem is solved by *t*_*s*_ of the form of Case 2. In fact, we have the following proposition.

#### Proposition 1

*Assume that the lockdown initiation time t*_*s*_ *is such that two relative maxima of I exist. Then the second maxima, occurring at some time t* ∈ [*t*_*s*_ + *T*, ∞), *is given by*

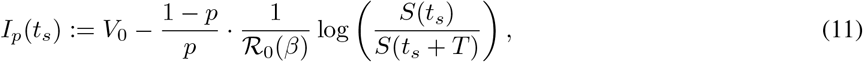

*where*

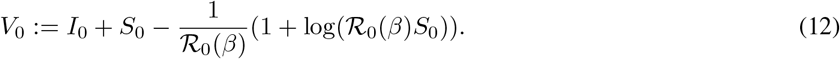

*Furthermore, V*_0_ *is the maximum of I corresponding to the case of a unique global maximum occurring in* [0, *t*_*s*_] *(Case 1 above). Thus, the minimization problem* (5) *is solved for t*_*s*_ *such that I admits two relative maxima (Case 2 above)*.

Recall that we are assuming that 0 < *p* < 1, which implies *S*(*t*_*s*_ + *T*) < *S*(*t*_*s*_). Since *I*(*t*_*s*_) ≤ *V*_0_ and the second term on the right-hand side of (11) is positive, we have that (5) is solved for *t*_*s*_ corresponding to Case 2 above, as claimed.

Before undertaking an analysis of *I*_*p*_(*t*_*s*_), we note that there are several equivalent representations of the second peak *I*_*p*_. By the change of variables *S* = *S*(*t*), we see that

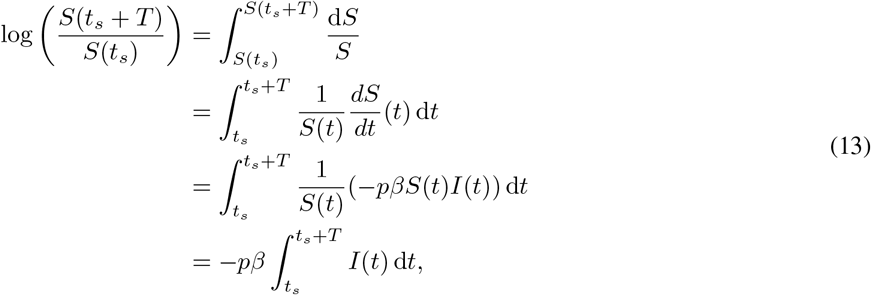

since 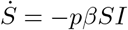 on (*t*_*s*_, *t*_*s*_ + *T*). Hence we can write (11) in the following form:

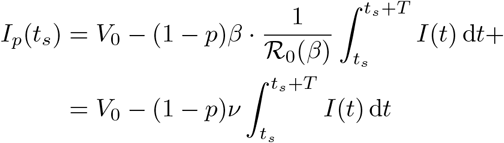

Since 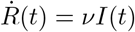, we can also write the previous integral as

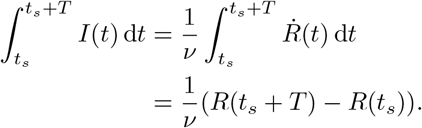

Thus, another equivalent form for *I*_*p*_(*t*_*s*_) is given by

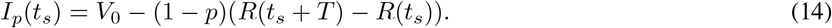

### 3.2 Analysis of relative maxima of *I*

Under assumptions (8), Proposition 1 implies that to minimize the peak of the infected population with respect to a lockdown represented by (4), we must minimize both relative maxima *I*(*t*_*s*_) and *I*_*p*_(*t*_*s*_) *simultaneously with respect to t*_*s*_. Recall that *I*(*t*_*s*_) denotes the infected population at the onset of the lockdown (i.e. *I* at time *t*_*s*_), and *I*_*p*_(*t*_*s*_) is the relative maxima of *I* occurring at some *t* ∈ [*t*_*s*_ + *T*, ∞), i.e. after the lockdown has been lifted. Specifically we note that we do not have an explicit formula for the time *I*_*p*_ occurs, and the notation is meant to emphasize that *I*_*p*_ depends on *t*_*s*_ via the expression (11).

Numerical simulations for a specific set of *β, p, ν, S*_0_ and *I*_0_ are provided in Figure 2. Here we simply vary the start time *t*_*s*_ for the distancing protocol represented by (4), and plot representative infection response dynamics in Figure 2a. Note that if the distancing starts too early (e.g. *t*_*s*_ = 20 days), then the peak of the infected population *I*_max_ is simply delayed until after the lockdown is lifted; *V*_0_ ≈ 0.4037 for the set of parameters in Figure 2. Similarly, if the lockdown is initiated too late (e.g. *t*_*s*_ = 70 days), then the distancing mandate has only a marginal effect on reducing *I*_max_. For intermediate *t*_*s*_, we observe two relative maxima *I*(*t*_*s*_) and *I*_*p*_(*t*_*s*_), as discussed in the beginning of this section. It appears that as *t*_*s*_ is increased, the first peak *I*(*t*_*s*_) increases, while the second peak *I*_*p*_(*t*_*s*_) decreases. This is intuitive, since increasing *t*_*s*_ allows us to initiate the lockdown closer to *V*_0_ (increasing *I*(*t*_*s*_)), which at the same time builds immunity in the population and hence decreases the magnitude of the “second wave” (which is quantified by *I*_*p*_(*t*_*s*_)). Furthermore, it appears that the optimal choice of *t*_*s*_ balances these two effects precisely:

**Figure 2:**
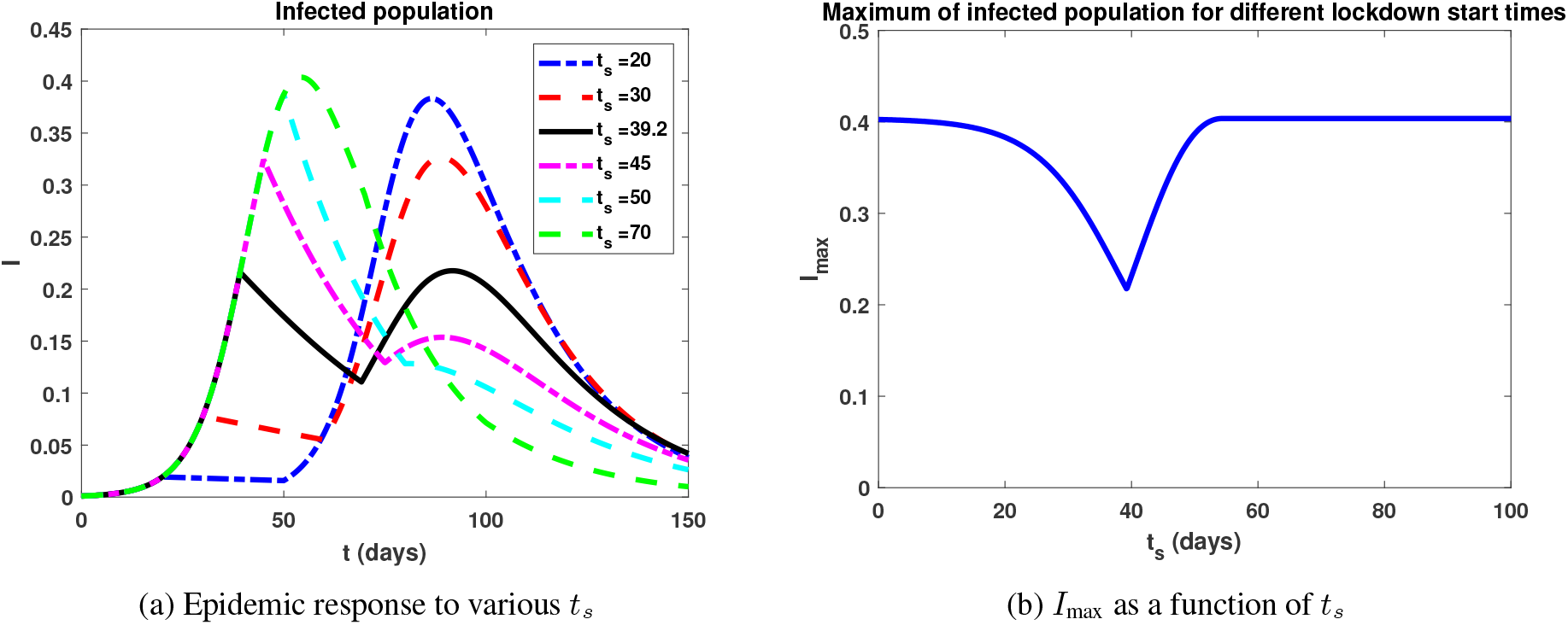
Response of infected population to a single lockdown as characterized by (4). The following parameters were utilized in the above simulation: *β* = 0.2, *p* = 0.2252, *ν* = 0.05, *I*_0_ = 10^−4^, and *S*_0_ = 1 − *I*_0_. Note that according to (7), ℛ_0_ = 4 prior to lockdown, while ℛ_0_ ≈0.9 during the lockdown. We observe a global minimum of *I*_max_ at *t*_*s*_ ≈39.2 days, which is corresponds to the black curve in Figure 2a. Note that both relative maxima appear to be equal for this minimizing *t*_*s*_.

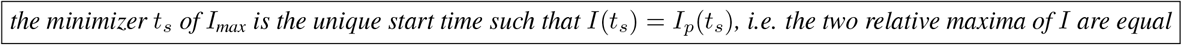

This hypothesis is generally true, and is stated precisely in the following theorem.

#### Theorem 1

*Consider the SIR epidemic model* (1) *with time-varying transmission rate* (4) *representing a single lock-down of relative efficacy p and fixed duration T* . *Under assumptions* (8), *the optimal time t*_*s*_ *of lockdown initiation to minimize the peak of the infected population is such that I has two relative maxima of equal size. In this case, the two maxima of I are precisely I*(*t*_*s*_) *and I*_*p*_(*t*_*s*_).

A proof of Theorem 1 is postponed until Section 5. Theorem 1 thus says that the optimal choice of *t*_*s*_ to “flatten the curve” is such that the infected population will have precisely two relative maxima of equal size: one of which occurs at lockdown initiation (*t*_*s*_), and the other given by a second wave occurring after the lockdown has been released (after *t*_*s*_ + *T*). We call this an *equal peaks* phenomena, with an unavoidable second wave of infections which will rebound to the same maximum intensity as experienced prior to the lockdown. We note that such a phenomenon occurs only for optimally designed interventions (indeed, it is a characterization), and it is possible to choose *t*_*s*_ large enough to reduce the second peak, or even remove it entirely (see the *t*_*s*_ = 70 days curve in Figure 2a). However, such distancing mandates will necessarily lead to a larger first peak in infections. Similarly, the first peak can be made as small as *I*_0_ if *t*_*s*_ is initiated early, but in this case a large second wave is encountered (see the *t*_*s*_ = 20 days curve in Figure 2a).

The result of Theorem 1 provides an implicit method of determining the optimal lockdown initiation time given the current state of the epidemic. The initiation time *t*_*s*_ is characterized by the relation

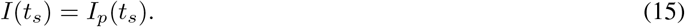

Utilizing (14) together with the (normalized conservation law)

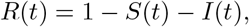

equation (15) can be written as

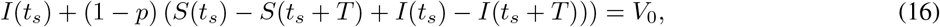

which yields an implicit relation to determine *t*_*s*_. Note that the time *t*_*s*_ cannot be computed explicitly (unlike the formulas which appear in [7]), since when *p >* 0 there are no analytic solutions to the SIR system in [*t*_*s*_, *t*_*s*_+*T*]. But of course, as the epidemic evolves, the relation (16) can be tested numerically. More precisely, policy could be designed by utilizing current epidemic data, assuming *t*_*s*_ = 0, and checking whether relation (16) is currently satisfied (with uncertainty sufficiently quantified, and assuming good estimates for lockdown efficacy *p* exist). If this equation is satisfied, the lockdown should be initiated as soon as possible. We finally note that other relations similar to (16) exist, which utilize the alternate forms of *I*_*p*_ presented in Section 3.1, as well as the conserved quantity *H*(*S, I*) to compute *I*(*t*_*s*_) as a function of *S*(*t*_*s*_) and the initial data (see the proof of Proposition 1 in Section 5). Specifically, a relation only involving *S*(*t*_*s*_) and *S*(*t*_*s*_ + *T*) is given as follows:

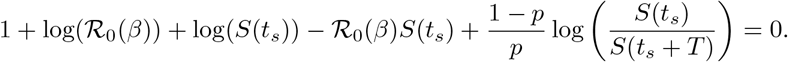

## 4 Discussion and extensions

In this work we have proven a characterization of the optimal start time with respect to (5) for a lockdown of fixed length *T* and transmission reduction factor *p* in the SIR model. This characterization is classified according to an *equal peaks* phenomenon: the infection response will exhibit two local maxima of equal magnitude. The result (Theorem 1) was proven under certain assumptions (8), but these were assumed for clarity of exposition, and the result remains valid under weaker hypothesis. In particular, if the lockdown is not “strong enough” (i.e. *p* does not satisfy the first of (8)), then the first relative maxima of *I* may occur interior to the lockdown interval [*t*_*s*_, *t*_*s*_ +*T*], and not at *t*_*s*_. But the result of Theorem 1 remains true: the optimal initiation time is such that both relative maxima are equal, i.e. the response of *I* possesses *equal peaks*.

The results of this work are concerned with optimizing the initiation time of a single *non-perfect* lockdown. In reality, social distancing directives are not designed utilizing a single interval of distancing, but more generally consist of multiple periods of possibly different levels of mandated distancing, which in the above model, correspond to different transmission reduction factors *p. We conjecture that the above equal peaks phenomenon generalizes to the case of multiple lockdowns*. More precisely, for *k* = 1, 2, …, *n* define *disjoint* intervals

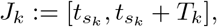

and *β*(*t*) be the time-varying transmission rate

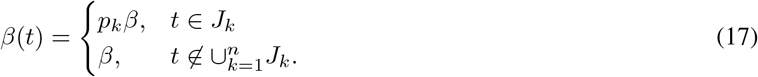

Here *β*(*t*) represents a series of *n* lockdowns, each of fixed (but generally different) length *T*_*k*_ and transmission reduction factor *p*_*k*_. A visualization for *n* = 2 is provided in Figure 3. We then can consider an optimization problem analogous to (5), where we minimize the peak of the infected population with respect to the start times 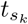 of the *k* = 1, 2, …, *n* lockdowns (assuming all other parameters are fixed and known). The conjecture generalizing Theorem 1 is then that the initiation times are such that the infected population exhibits *n* + 1 relative maxima, each of equal size; that is, it possesses *n* + 1 equal peaks. Although we do not prove the result here, numerical simulations seem to suggest its validity. A numerical experiment for *n* = 2 is provided in Figure 4a. Here we simply iterate over all possible intervals 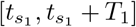 and 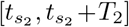 that do not overlap, as we cannot start a second lockdown before the first one has ended; these prohibited times correspond to the white region in Figure 4a, and is given parametrically by 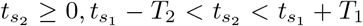 in the first quadrant. Bluer shades in Figure 4a correspond to smaller infection peaks, and the approximate optimal policy together with the infection response is provided in Figure 4b. Note that the infected population exhibits 3 = *n*+1 relatives maxima (peaks) of equal size, as hypothesized.

**Figure 3:**
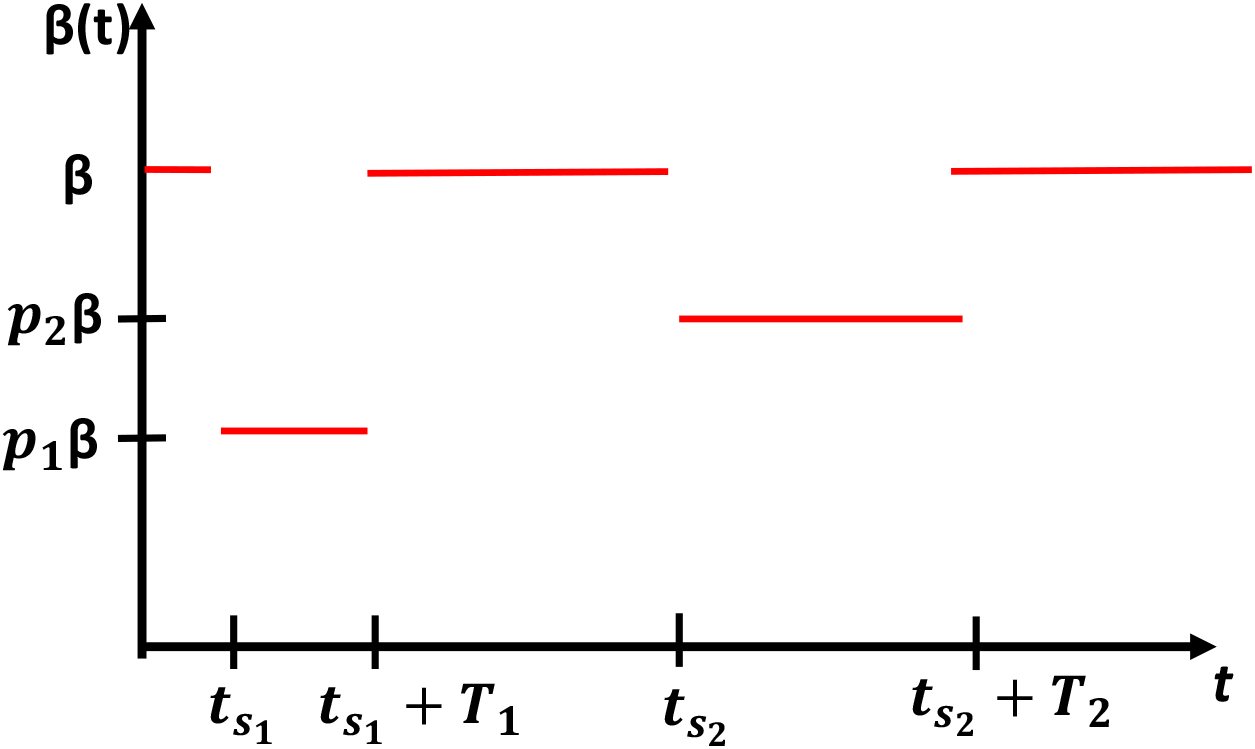
Visualization of *n* = 2 lockdowns (17). We assume that transmission is reduced by an effective amount *p*_1_ during 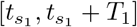 and *p*_2_ during 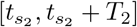, where 0 < *p*_1_, *p*_2_ < 1.

**Figure 4:**
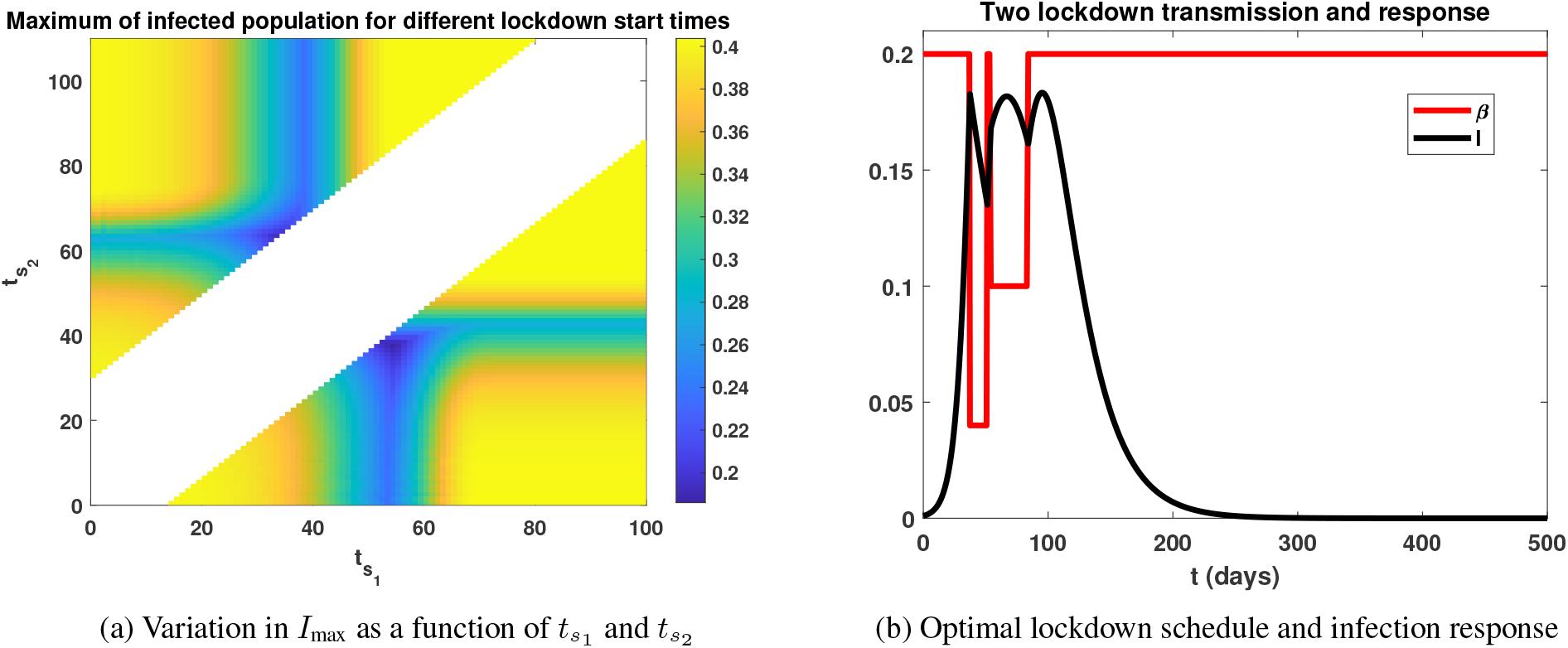
Numerical investigation of optimal lockdown schedule of the form (17) with *n* = 2. Parameters utilized are *β* = 0.2, *p*_1_ = 0.5, *p*_2_ = 0.2, *T*_1_ = 30, *T*_2_ = 14, *ν* = 0.05, *I*_0_ = 10^−4^, and *S*_0_ = 1 − *I*_0_. As in Figure 2, our goal is to minimize the peak of the infected population, which corresponds to bluer shades in Figure 4a. The white region is prohibited, as it would correspond to overalapping lockdowns. The optimal initiation times 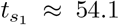 days and 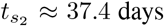 days is provided in Figure 4b. Note that the response *I* exhibits 3 = *n* + 1 relative maxima (peaks) of equal size.

## 5 Proofs of results

In this section, we provide proofs of all results stated in the above manuscript, including our main result Theorem 1. We also present an intermediate proposition (Proposition blah) which allows us to compute the sensitivities of the susceptible population (and also the infected population) with respect to the initiation time *t*_*s*_; this is the main tool used to prove Theorem 1.

*Proof of Proposition 1*. The following calculation is tedious, but elementary. It is well known that the quantity

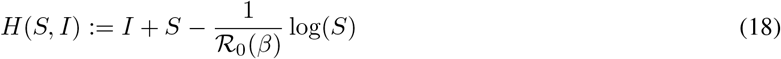

is conserved along solutions curves with constant *β* and *ν*. Since transmission rates of the form (4) are piecewise constant, we thus need to consider two two transmission rates: *β* and *pβ*. The second local maximum of *I* must occur after the lockdown is released, since *İ* ≤ 0 on (*t*_*s*_, *t*_*s*_ + *T*). We use repeated use of the conserved quantity *H*(*S, I*) on intervals where *β*(*t*) is constant. Since *I* is guaranteed to increase after the lockdown is released (assumption (9)), *t*_*s*_ + *T* is a local minimum of *I*, and hence the local maximum must occur on the open interval (*t*_*s*_ + *T*, ∞). On this interval, *β*(*t*) ≡ *β* (i.e. ℛ_0_ is given by ℛ_0_(*β*)), we have the conservation law

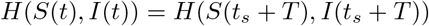

for *t* ≥ *t*_*s*_ + *T* . Thus, we can solve for *I* as a function of *S* and the initial conditions (*S*(*t*_*s*_ + *T*), *I*(*t*_*s*_ + *T*)) along trajectories:

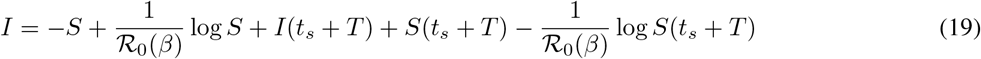

A relative maximum of *I* with respect to *t* thus corresponds to a relative maximum of *I* with respect to *S*, which must occur at a susceptible population *S*_∗_ such that (*dI/dS*)(*S*_∗_) = 0. Utilizing (19) above then implies that *S*_∗_ is given by

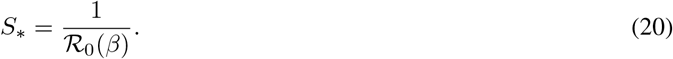

Note that *S*_∗_ corresponds to the herd immunity population for the SIR model, and (9) ensures that *S*_∗_ = *S*(*t*_∗_) satisfies

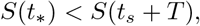

i.e. that

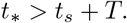

Substituting (20) into (19) yields an expression for the second peak *I*_*p*_ of *I*:

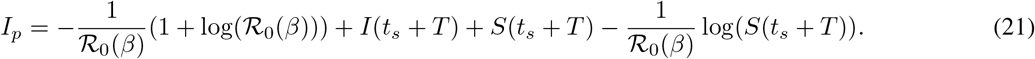

We now iterate the above procedure on [*t*_*s*_, *t*_*s*_ + *T*] to simplify the above expression for *I*_*p*_ (by simplify, we mean remove as many dependencies on time *t*_*s*_ + *T* as possible). On [*t*_*s*_, *t*_*s*_ + *T*], *β*(*t*) ≡ *pβ*, and the conservation law implies that

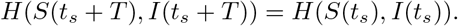

Thus, we can solve for *I*(*t*_*s*_ + *T*) + *S*(*t*_*s*_ + *T*) as

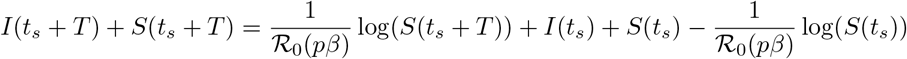

Substituting this into (21) yields

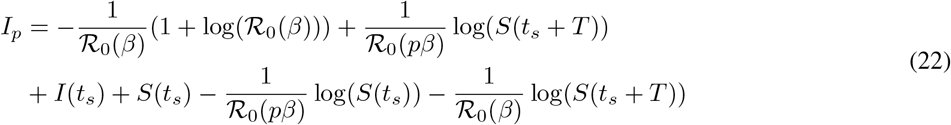

Lastly on [0, *t*_*s*_], where *β*(*t*) ≡ *β*, we have that

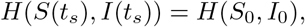

which allows us to solve for *I*(*t*_*s*_) + *S*(*t*_*s*_) as

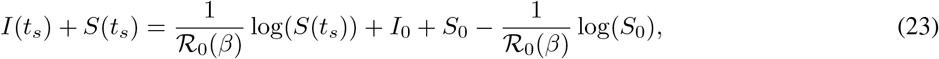

which yields in (22) the expression

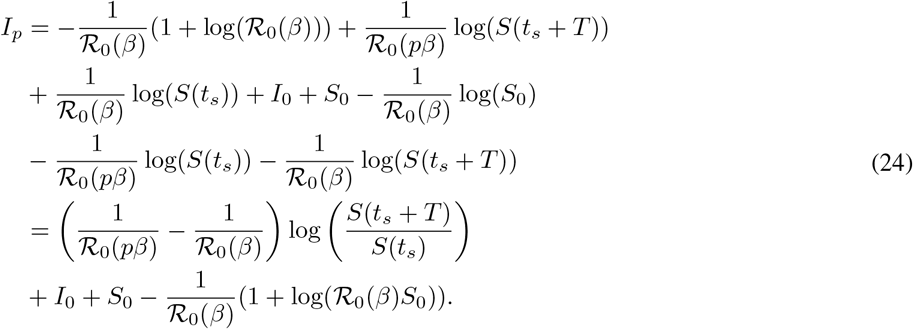

We can further simplify (24) by noting that

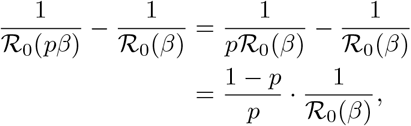

and defining the constant *V*_0_ as in (12), we have that the second relative maximum of *I, I*_*p*_ = *I*_*p*_(*t*_*s*_), is given by

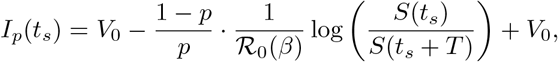

as claimed.

We note that *V*_0_ is the maximum value of *I* if a lockdown is never implemented (*β*(*t*) ≡ *β* on [0, ∞)), or if the lockdown is “too late” to reduce the peak of the infected population. Equivalently, a peak of *V*_0_ corresponds to a locktown initiation *t*_*s*_ such that

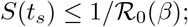

compare this to (10). In such cases, *I* has only one (global) maximum *V*_0_, and is a subset of Case 1 in Section 3.1 (the other disjoint subset is when the lockdown length *T* is long enough so that the global maximum of *I* occurs at *t*_*s*_, and no other local maxima exist). □

Before stating and proving the next proposition, we note that the conserved quantity (18) allows us to transform the two-dimensional system (1) into a one-dimensional (nonlinear) ODE *if β*(*t*) *is constant*. Specifically, it allows us to solve for *I*(*t*) as a function of *S*(*t*). For example, on [0, *t*_*s*_] where *β*(*t*) ≡ *β*, we have that

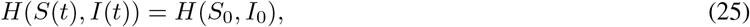

or equivalently,

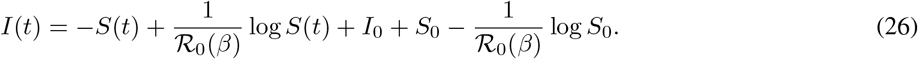

Thus, the dynamics on [0, *t*_*s*_] can be understood by analyzing the one-dimensional ODE

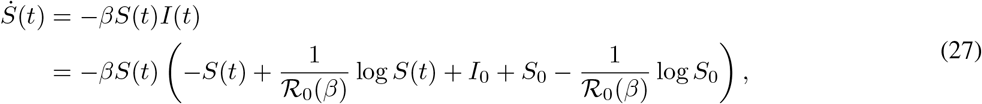

with the infected population given by (26). Similarly, on [*t*_*s*_, *t*_*s*_ + *T*],

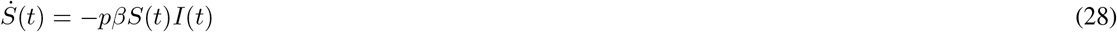

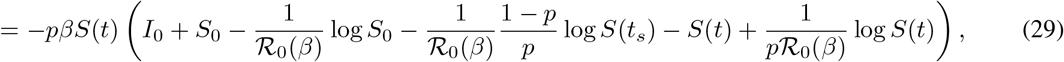

where we have used (23) to replace *I*(*t*_*s*_) + *S*(*t*_*s*_).

### Proposition 2

*For any fixed t >* 0, *S* = *S*(*t*; *t*_*s*_) *is differentiable with respect to the lockdown initiation time t*_*s*_; *call this derivative the sensitivity of S with respect to t*_*s*_ *at time t. Furthermore, we have the following formulas for the sensitivities of S at times t*_*s*_ *and t*_*s*_ + *T* :

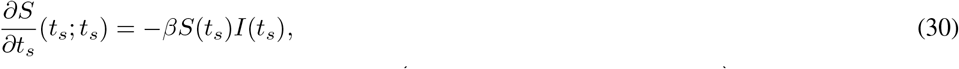

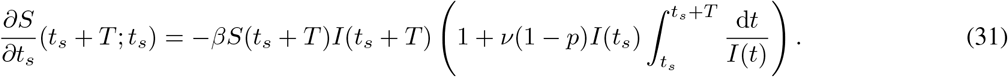

*Proof of Proposition 2*. Differentiability is clear. In the following, we suppress the dependence of *S* on the parameter *t*_*s*_ for notational ease. To see (30), we utilize (27), which we write in the form

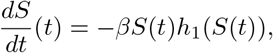

with 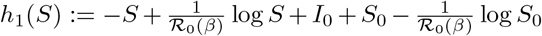. Note by the argument preceding the proposition we have that *h*_1_(*S*(*t*)) = *I*(*S*(*t*)) (see (27)). Since the above is autonomous and one-dimensional, it is separable, and hence we have the relation

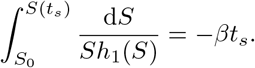

Differentiating with respect to *t*_*s*_ yields the equation

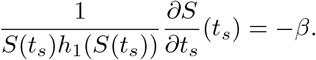

Thus,

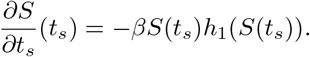

As mentioned previously, *h*_1_(*S*(*t*_*s*_)) = *I*(*t*_*s*_), which yields (30).

We perform the same calculation to obtain (31). That is, write

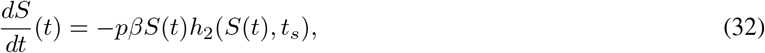

on [*t*_*s*_, *t*_*s*_ + *T*], with

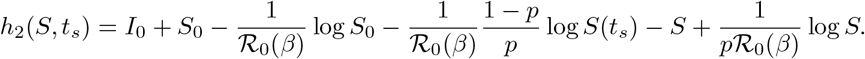

Note that in this case, *h*_2_ has an additional *t*_*s*_ dependence via the *S*(*t*_*s*_) term. Separating and integrating (32) yields the relation

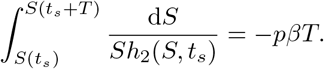

Differentiating the above (using the Leibniz rule, since *t*_*s*_ appears in both the bounds and the integrand), we obtain

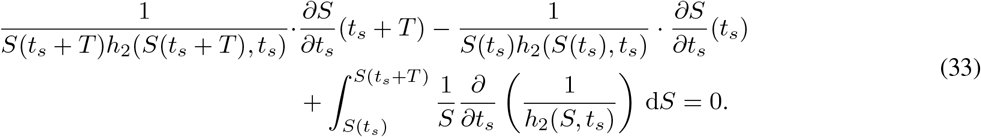

The integral term can simplified as follows:

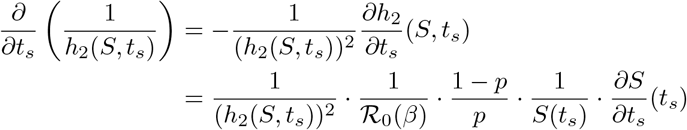

Hence

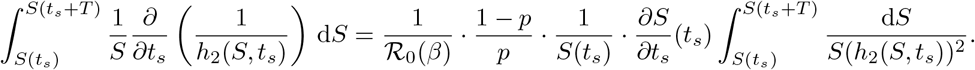

Recalling that *h*_2_(*S, t*_*s*_) = *I* on [*t*_*s*_, *t*_*s*_ + *T*] and changing variables on the integral (*S* = *S*(*t*), as in the first line of (13)), we obtain

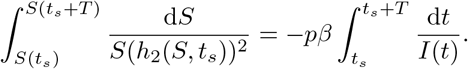

Thus the third term on the left-hand side of (33) becomes

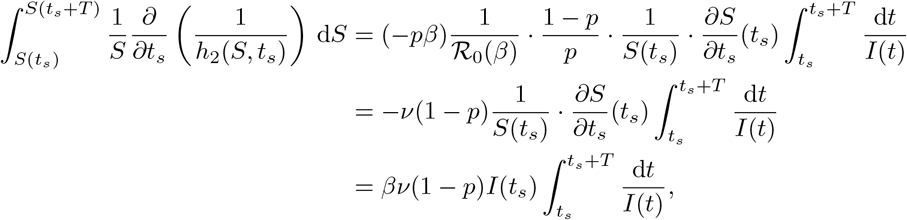

where in the final equality we used (30). We can now solve (33) for (*∂S/∂t*_*s*_)(*t*_*s*_ + *T*), again using (30) and the fact that *h*_2_(*S*(*t*_*s*_ + *T, t*_*s*_) = *I*(*t*_*s*_ + *T*), *h*_2_(*S*(*t*_*s*_), *t*_*s*_) = *I*(*t*_*s*_) to obtain

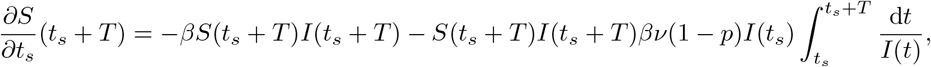

□

which is precisely (31).

*Proof of Theorem 1*. We have already seen that *I* must have two relative maxima for *I*_max_ to be minimized, and we have also seen that these maxima are given by *I*(*t*_*s*_) and *I*_*p*_(*t*_*s*_). Thus, all that remains to show is that the minimizing *t*_*s*_ of (5) must occur when *I*(*t*_*s*_) = *I*_*p*_(*t*_*s*_). Note that it will be sufficient to show that *I*(*t*_*s*_) increases as a function of *t*_*s*_ (again, assuming (8)), and that *I*_*p*_(*t*_*s*_) decreases as a function of *t*_*s*_, and that they have a (thus necessarily unique) point of intersection. Note that the latter is clear, assuming *I*_0_ is small, since *I*(*t*_*s*_) →*I*_0_ and *I*_*p*_(*t*_*s*_) →*V*_0_ as *t*_*s*_ →0. Hence all that remains is to show the monotonicity properties of *I*(*t*_*s*_) and *I*_*p*_(*t*_*s*_).

Note that is intuitively clear that *I*(*t*_*s*_), the value of the first relative maxima of *I*, increases as a function of *t*_*s*_, since (*dI/dt*)^−^(*t*_*s*_) *>* 0 by (10), so increasing *t*_*s*_ increases *I*(*t*_*s*_). More rigorously, via the conserved quantity *H* (see equation (18)), we have that

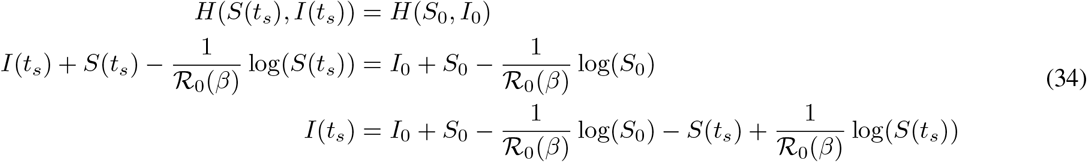

Taking a derivative with respect to *t*_*s*_ of the latter equality yields

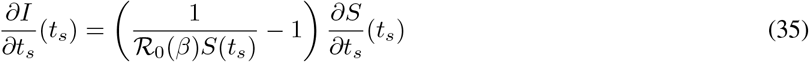

Since ℛ_0_(*β*)*S*(*t*_*s*_) *>* 1 (this is again (10), which is a consequence of *I* possessing a local maximum at *t*_*s*_), it is sufficient to show that *S*(*t*_*s*_) decreases as a function of *t*_*s*_ at *t* = *t*_*s*_. But this is clear from (30) of Proposition 2.

To prove that *I*_*p*_(*t*_*s*_) decreases as a function of *t*_*s*_, note that by (11), it is sufficient to prove that *S*(*t*_*s*_)*/S*(*t*_*s*_ +*T*) is increasing as a function of *t*_*s*_, or equivalently that *S*(*t*_*s*_ + *T*)*/S*(*t*_*s*_) is decreasing as a function of *t*_*s*_. Since *S* is differentiable with respect to *t*_*s*_ for all fixed *t*, it is sufficient to analyze the sign of the derivative of *S*(*t*_*s*_ + *T*)*/S*(*t*_*s*_). By elementary calculus, we have that

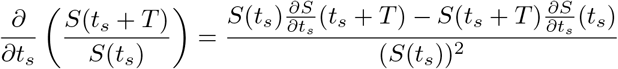

We can now use formulas (30) and (31) in Proposition 2 to compute the numerator:

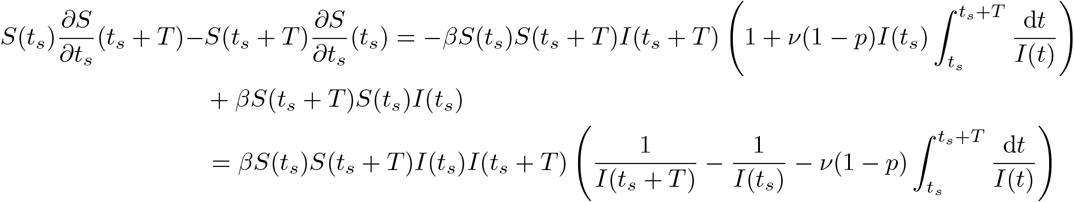

To complete the proof, we thus need to show that the parenthetical term on the right-hand side of the above is negative. Equivalently, we need to show that

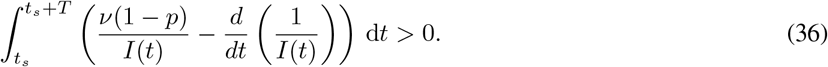

On (*t*_*s*_, *t*_*s*_ + *T*), we have that (since *İ* = *pβSI* − *νI*)

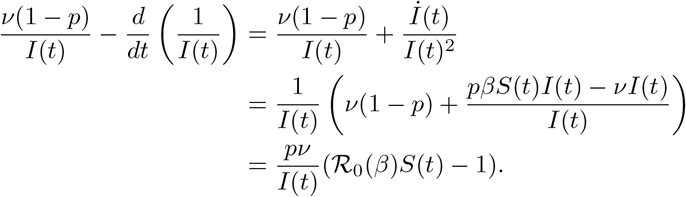

As *S* is non-increasing, we have that *S*(*t*) ≥ *S*(*t*_*s*_ + *T*) for all *t* ∈ (*t*_*s*_, *t*_*s*_ + *T*). This together with assumption (9) imply that

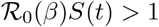

for all *t* ∈ (*t*_*s*_, *t*_*s*_ + *T*). Hence the integrand in (36) is positive, which is sufficient to complete the proof. □

## Data Availability

N/A

## Data Availability

N/A

